# Scoping review protocol- Scoping review Vitamin B_12_ deficiency in long term metformin use and clinician awareness

**DOI:** 10.1101/2025.02.17.25322407

**Authors:** Ian Parsonage, David Wainwright, Julian Barratt

## Abstract

**Introduction:** A relationship between long-term metformin use and vitamin B_12_ deficiency has been long discussed in literature. Nonetheless, prior to 2022, there was no official guidance. In June 2022, the MHRA published advice, stating that low vitamin B_12_ is now considered to be a common side effect. It advises checking levels in patients with symptoms of B_12_ deficiency, as well as monitoring those at risk of B_12_ deficiency.

Despite efforts to promote evidence-based practice, there is still a gap in the translation of research findings into policies and clinical practice. The above research has been shared widely in the academic and specialist Diabetes literature over a prolonged period. The purpose of the scoping review is to explore what evidence is available regarding clinicians’ awareness of the association between metformin use and vitamin B_12_ deficiency in patients with T2DM and how this evidence is implemented into frontline clinical practice and what screening processes are recommended or exist?

**Methods and analysis:** This is a protocol for a scoping review to be guided by the Joanna Briggs Institute (JBI) methodology for scoping reviews ^20^. The databases to be searched will include CINAHL, MEDLINE, EMBASE, British Nursing Index, via EBSCOhost. Sources of unpublished studies, policies and grey literature will include Google Scholar, the Cochrane Library, ProQuest Dissertations and Theses Open. Titles and abstracts of articles were reviewed by the authors. If articles were representative of the inclusion criteria, the articles went through a full-text review by the authors The results of the search and study inclusion/exclusion process will be reported and presented in a Preferred Reporting Items for Systematic Reviews and Meta-analyses (PRISMA) flow diagram. Data will be extracted from papers, using the recommended JBI data extraction tool.

**Strengths and limitations of this study:**

- To the authors’ knowledge, this is the first scoping review to look at clinician awareness of Vitamin B_12_ deficiency in long term metformin use.
- Strengths include an extensive search of multiple databases, including grey literature.
- As this is a scoping review, the quality of the evidence and risk of bias will not be evaluated.
- The review is also limited to selected languages which may bias the evidence.

## Introduction

Diabetes is a serious and growing global health concern, according to the latest statistics. The global prevalence of impaired glucose tolerance was estimated at 7.5% (374 million) in 2019 and is projected to reach 8.0% (454 million) by 2030 and 8.6% (548 million) by 2045 ^1^. The leading drug in the treatment of diabetes is metformin hydrochloride, with approximately 24.1 million items dispensed ^2^ with one study showing 83.6% of Type 2 Diabetes Mellitus (T2DM) patients take metformin ^3^.

All guidelines, including the European Association for the Study of Diabetes (EASD) and the American Diabetes Association (ADA), consider metformin a cornerstone and first-line treatment, along with lifestyle intervention, for managing hyperglycaemia in patients with Type 2 Diabetes Mellitus (T2DM) ^4^. In the UK, metformin is the recommended first-line therapy for the treatment of T2DM in patients with normal renal function and is widely used ^5^.

As far back as 1971, this effect was being studied with Tomkin ^6^ reporting that approximately 30% of patients taking metformin do not properly absorb vitamin B_12_. A study published in 2010^7^ found that, compared with placebo, use of metformin 850□mg three times a day for 4 years was associated with a mean decrease in vitamin B_12_ concentration of almost 20%.

The first documented association were published in the late 1960s when annual serum vitamin B_12_ testing was already suggested as a valid screening measure for early detection of vitamin B_12_ deficiency in patients on long-term metformin therapy ^6,8^. However, definitive screening guidelines are lacking.

In 2021, the American Diabetes Association (ADA) Standards of Medical Care in Diabetes recommend considering a periodic assessment of vitamin B_12_ levels in patients with long-term metformin use, including those with prediabetes, peripheral neuropathy or anaemia ^9^. Infante undertook a ‘field of vision’ article into the link between Vitamin B_12_ and metformin and suggested screening guidelines based on the current evidence and propose a list of criteria for a cost-effective vitamin B_12_ deficiency screening in metformin-treated patients ^10^. Nonetheless, to date, no definite guidelines are available for the screening of vitamin B_12_ deficiency in patients taking metformin.

The known adverse drug reaction of vitamin B_12_ deficiency was recently reviewed for the brand leader Glucophage (metformin) within Europe with input from the MHRA ^11^. After this review, MHRA has agreed that the product information for medicines containing metformin should be updated.

In June 2022, the MHRA published advice, which concluded that this side effect occurs more frequently than was previously thought ^11^. This has led to an update to the Product Information for all metformin-containing medications. The MHRA states that low vitamin B_12_ is now considered to be a common side effect, especially when taking high-dose or long-term metformin, affecting up to one in 10 people ^11^. It advises checking levels in patients with symptoms of B_12_ deficiency, as well as monitoring those at risk of B_12_ deficiency ^11^.

Prior to an MHRA alert ^11^ there was no guidance issued on this matter in the UK ^12^. Currently, in the UK, the Clinical Knowledge Summary (CKS) lists vitamin B_12_ deficiency as an adverse effect ^13^.

Awareness of monitoring for adverse effects from medication has been shown to be poor in other similar cases. A recent supported this view and highlight how individual clinicians find it difficult to be aware of all the relevant, valid evidence, especially with many important practice changes linked to low-cost pharmaceuticals or non-pharmaceuticals ^14^.

Despite efforts to promote evidence-based practice, there is still a gap in the translation of research findings into policies and clinical practice ^15^. The National Institute for Health and Care research (NIHR) agrees and argues when such interventions are implemented (or put into practice), the process is often challenging, unpredictable and typically slow ^16^.

The research surrounding the risk of vitamin B_12_ deficiency with metformin use has been shared widely in the academic and specialist Diabetes literature over a prolonged period. However, a MHRA alert had to be released to highlight this risk to clinicians surrounding this adverse effect. The purpose of the scoping review is to explore what evidence is available regarding clinicians’ awareness of the association between metformin use and vitamin B_12_ deficiency in patients with T2DM and how this evidence is implemented into frontline clinical practice and what screening processes are recommended or exist?

Scoping reviews are now seen as a valid approach in those circumstances where systematic reviews are unable to meet the necessary objectives or requirements of knowledge users ^17^. There now exists clear guidance regarding the definition of scoping reviews, how to conduct scoping reviews and the steps involved in the scoping review process ^18^. A scoping review may be applicable if authors do not always wish to ask such single or precise questions and may be more interested in the identification of certain characteristics/concepts in papers or studies, and in the mapping, reporting or discussion of these characteristics/concepts. In these cases, a scoping review is the better choice ^19^.

A scoping review was considered a more optimal choice over a systematic review for the question posed. The question covers several concepts and themes in which individual studies and reviews exist, but no mapping has taken place exploring these themes together.

A preliminary search of MEDLINE, the Cochrane Database of Systematic Reviews and JBI Evidence Synthesis was conducted and no current or underway systematic reviews or scoping reviews on the topic were identified.

### Review question

1. **Clinician awareness**: What is the extent of evidence regarding clinician awareness of vitamin B12 deficiency as a side effect of long-term metformin use in patients with type 2 diabetes?
2. **Screening and monitoring practices**: What are the existing screening and monitoring practices in primary care settings for early detection of vitamin B12 deficiency in patients on long-term metformin?
3. **Implementation into clinical practice**: How is the evidence on clinician awareness and screening practices for vitamin B12 deficiency implemented in frontline clinical practice for patients with type 2 diabetes on long-term metformin?

## Methods and analysis

### Methods

The proposed scoping review will be based on the JBI (Joanna Briggs Institute) methodology for scoping reviews ^20^. Prior to this review, a thorough review of the literature will be undertaken. Systematic reviews may exist for each component of the title (vitamin B_12_ deficiency in patients on long term metformin, clinician awareness of evidence-based practice, screening/early detection), but the scoping review will continue if no previous scoping reviews or studies have been undertaken looking at screening methods for metformin related vitamin B_12_ deficiency or clinician awareness of vitamin B_12_ deficiency in patients on long term metformin use. Therefore, this scoping review will be used to identify and analyse knowledge gaps.

The structure of this scoping review will be based on the framework proposed by Arksey and O’Malley ^21^ and was revised by Peters ^17^(see figure 1). The methodological process will be based on the JBI approach. Preferred Reporting Items for Systematic reviews and Meta-Analyses extension for Scoping Reviews (PRISMA-ScR) will be used for reporting of the scoping review as per the BMJ guidelines ^22^.

**Figure 1:**
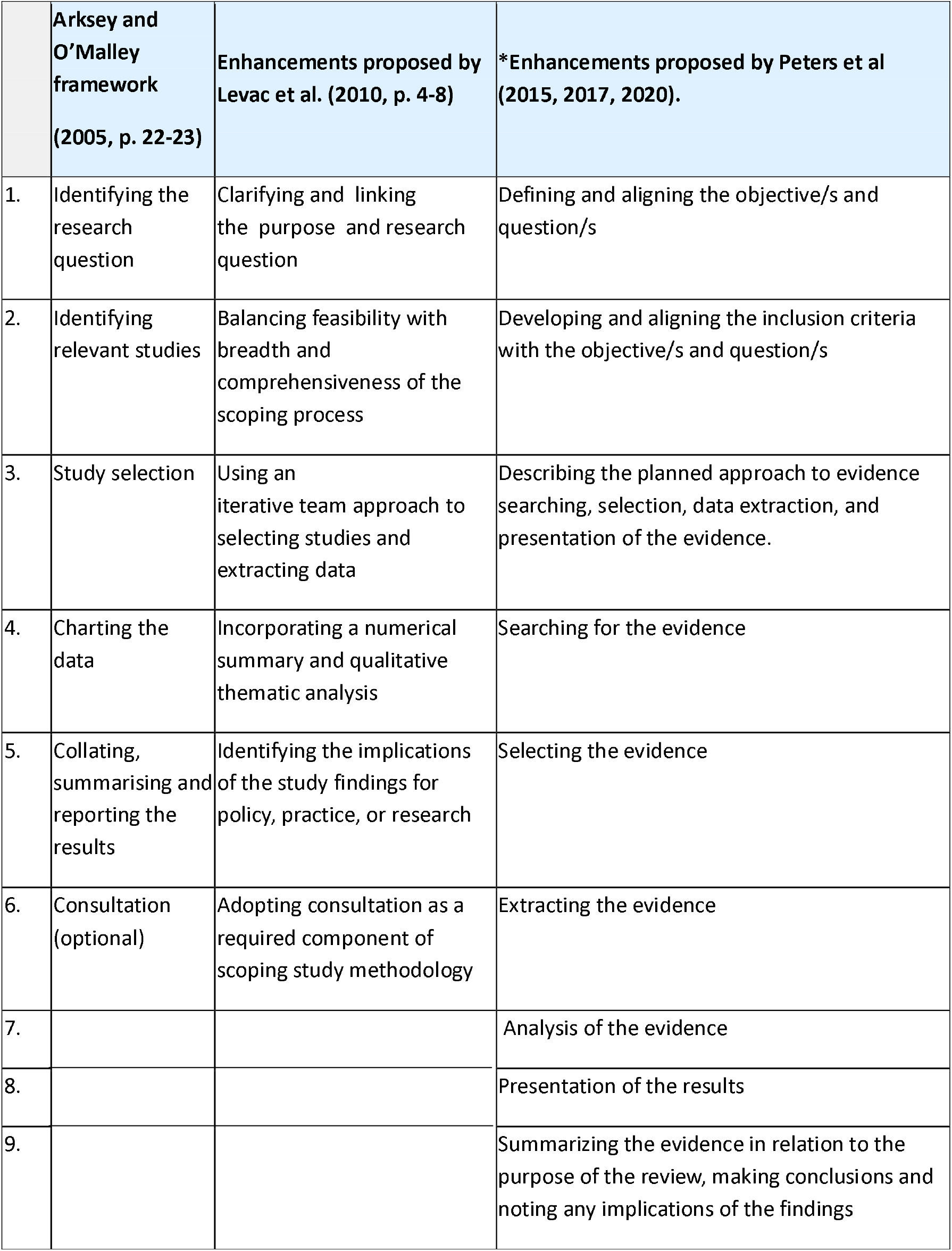
Framework

### Search strategy

The search strategy will aim to locate both published and unpublished studies. The text words contained in the titles and abstracts of relevant articles, and the index terms used to describe the articles will be used to develop a full search strategy.

The search strategy, including all identified keywords and index terms, will be adapted for each database and/or information source. The reference list of all included sources of evidence will be screened for additional studies.

Only studies published only in English will be included. (See figure 2 for search strategy).

**Figure 2:**
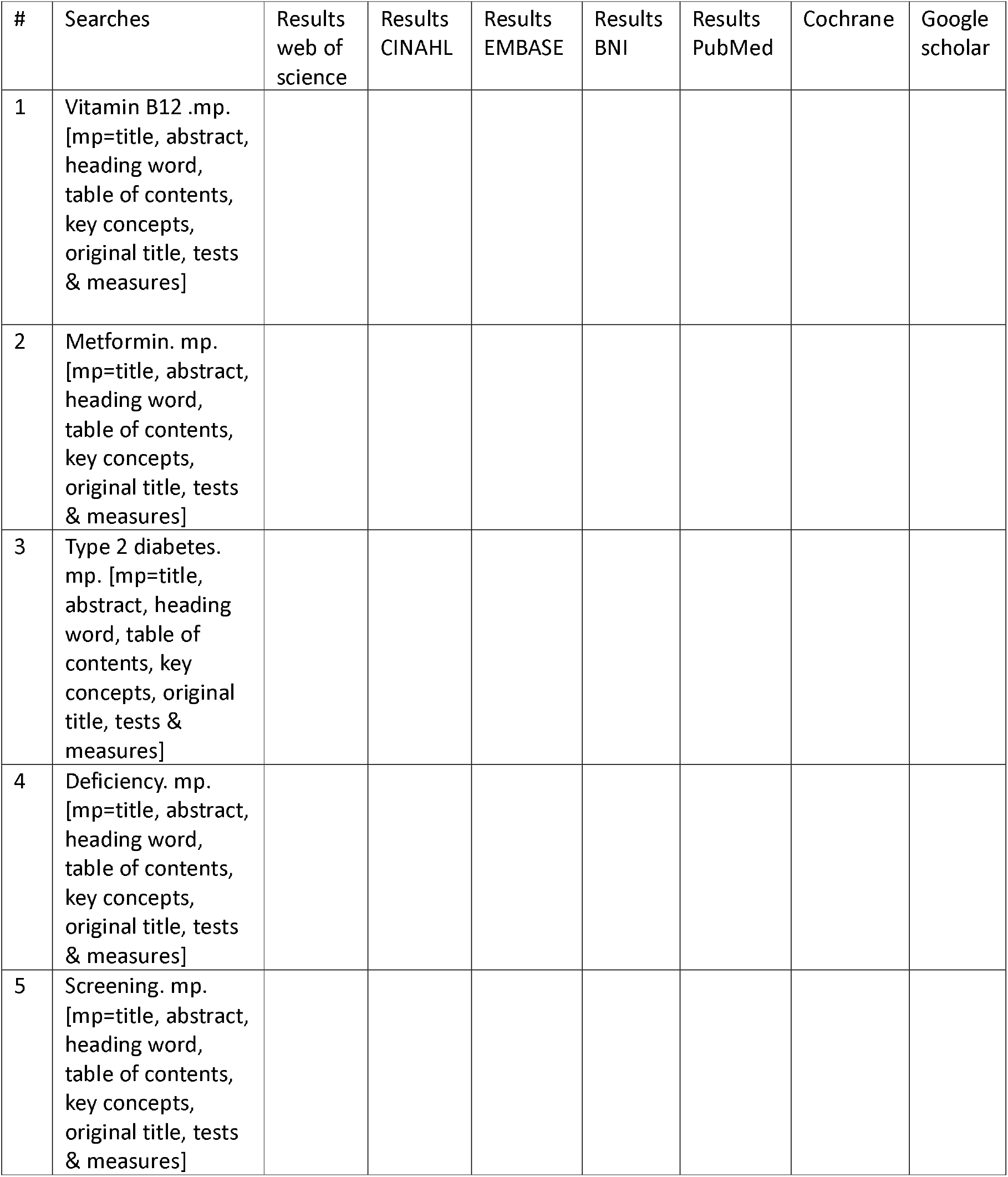
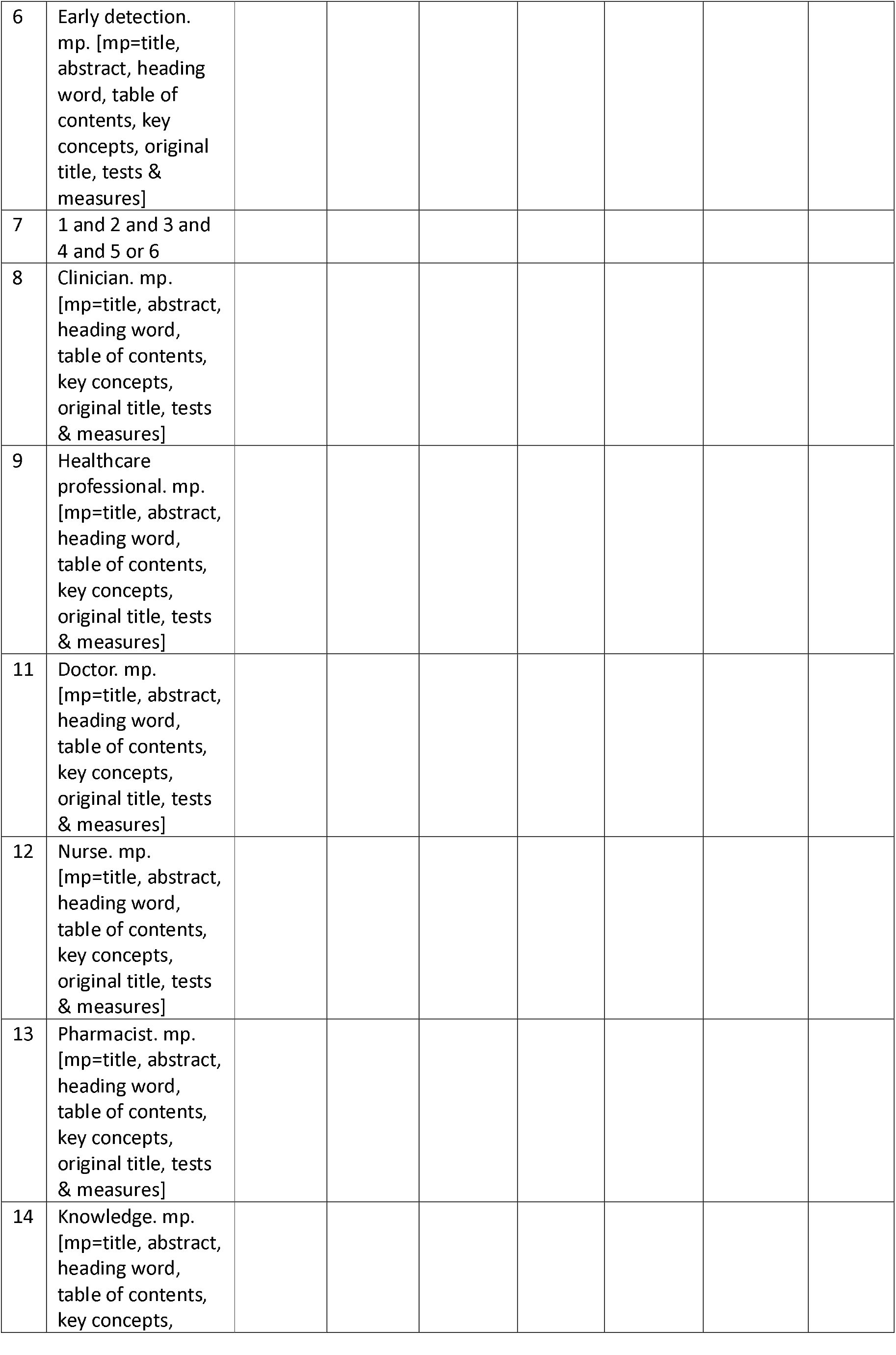
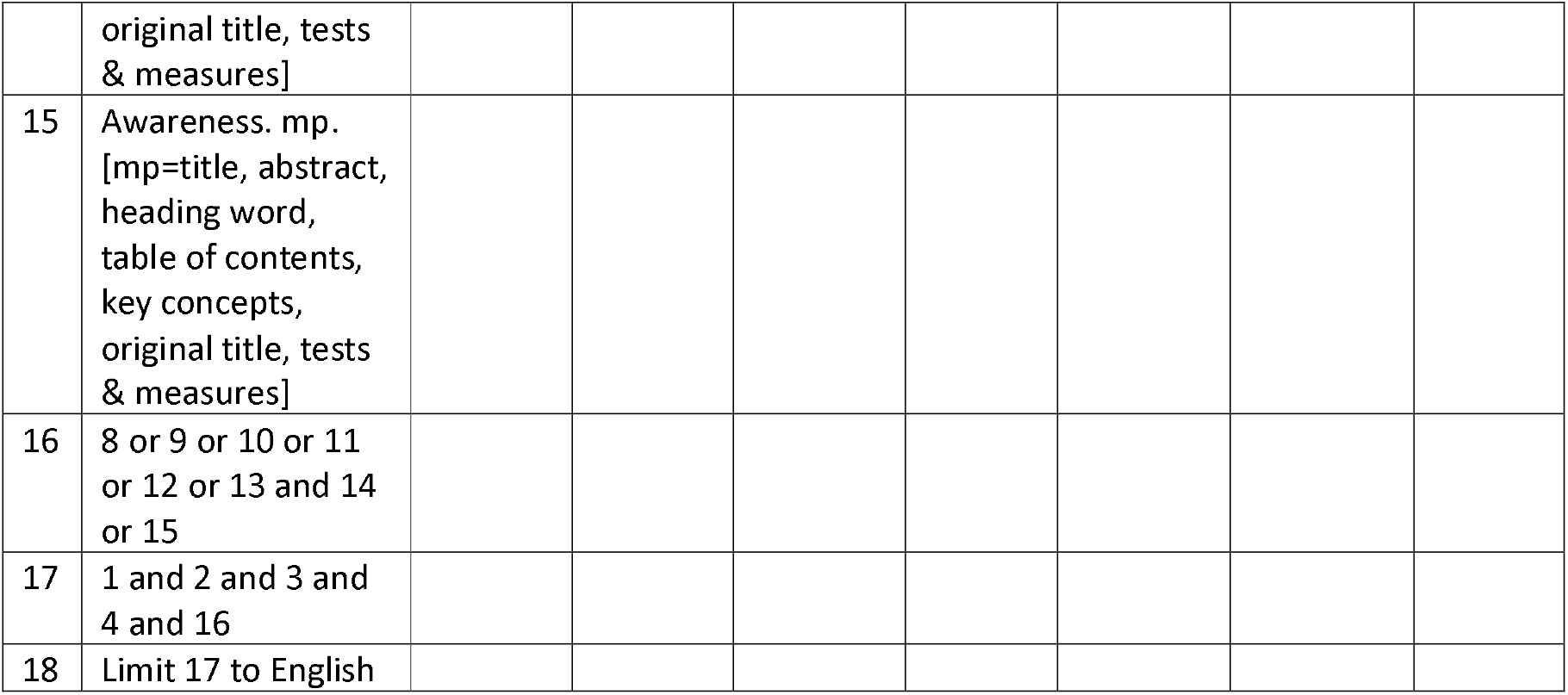
Search strategy example

The databases to be searched include MEDLINE (PubMed), British Nursing Index (BNI), Google scholar, Cochrane, Embase, Web of Science and CINAHL (EBSCO) alongside searching for grey literature such as EThOS, DART European and Kings College London Research Portal.

### Study/Source of evidence selection

Following the search, all identified citations will be collated and uploaded into EndNote 21 Clarivate Analytics, PA, USA. The full text of selected citations will be assessed in detail against the inclusion criteria. Only articles published since 1990 and written in English were eligible for inclusion in this review. Articles were excluded if they did not pertain to clinician awareness of or screening of, vitamin B12 deficiency in type 2 diabetes or metformin use. Titles and abstracts of articles were reviewed by the authors. If articles were representative of the inclusion criteria, the articles went through a full-text review by the authors. The results of the search and the study inclusion process will be reported in full in the final scoping review and presented in a PRISMA flow diagram ^23^.

### Inclusion criteria

#### Study Population

Participants over 18 with type 2 diabetes mellitus on long term metformin. Registered healthcare professionals who prescribe or monitor patients with Type 2 diabetes and on long term metformin.

#### Concept

Papers need to look at vitamin B_12_ deficiency linked to long term metformin use. Papers that explore clinician knowledge of side effects of metformin long term. Papers where the full text are immediately available. Primary care professionals who are implementing screening protocols/monitoring protocols strategies for vitamin B_12_ deficiency in relation to long term metformin use.

#### Context

Any healthcare settings (including international) in which registered healthcare professionals are responsible for the monitoring and prescribing of medication for patients with type 2 diabetes including metformin. These articles must be published after 1990.

#### Types of sources

This scoping review will consider both qualitative and quantitative studies including experimental and quasi-experimental study designs including randomized controlled trials, non-randomized controlled trials, before and after studies and interrupted time-series studies. In addition, analytical observational studies including prospective and retrospective cohort studies, case-control studies and analytical cross-sectional studies will be considered for inclusion. This review will also consider descriptive observational study designs including case series, individual case reports and descriptive cross-sectional studies for inclusion.

In addition, systematic reviews that meet the inclusion criteria will also be considered, depending on the research question.

#### Data extraction

Data will be extracted from papers included in the scoping review by the reviewer using a data extraction chart developed by the author (See figure 3). The data extraction was conducted by the lead author using a data extraction sheet that was developed after critical discussion. This sheet included information about the name of study, author, year of publication, origin/country, aims/purpose, population and sample size, methodology/methods, outcomes and details of these key findings that relate to scoping review. For extracting data from the discussion papers, the emphasis was placed on extracting apparent themes from the papers and then interpreting those themes using personal judgements.

**Figure 3:**
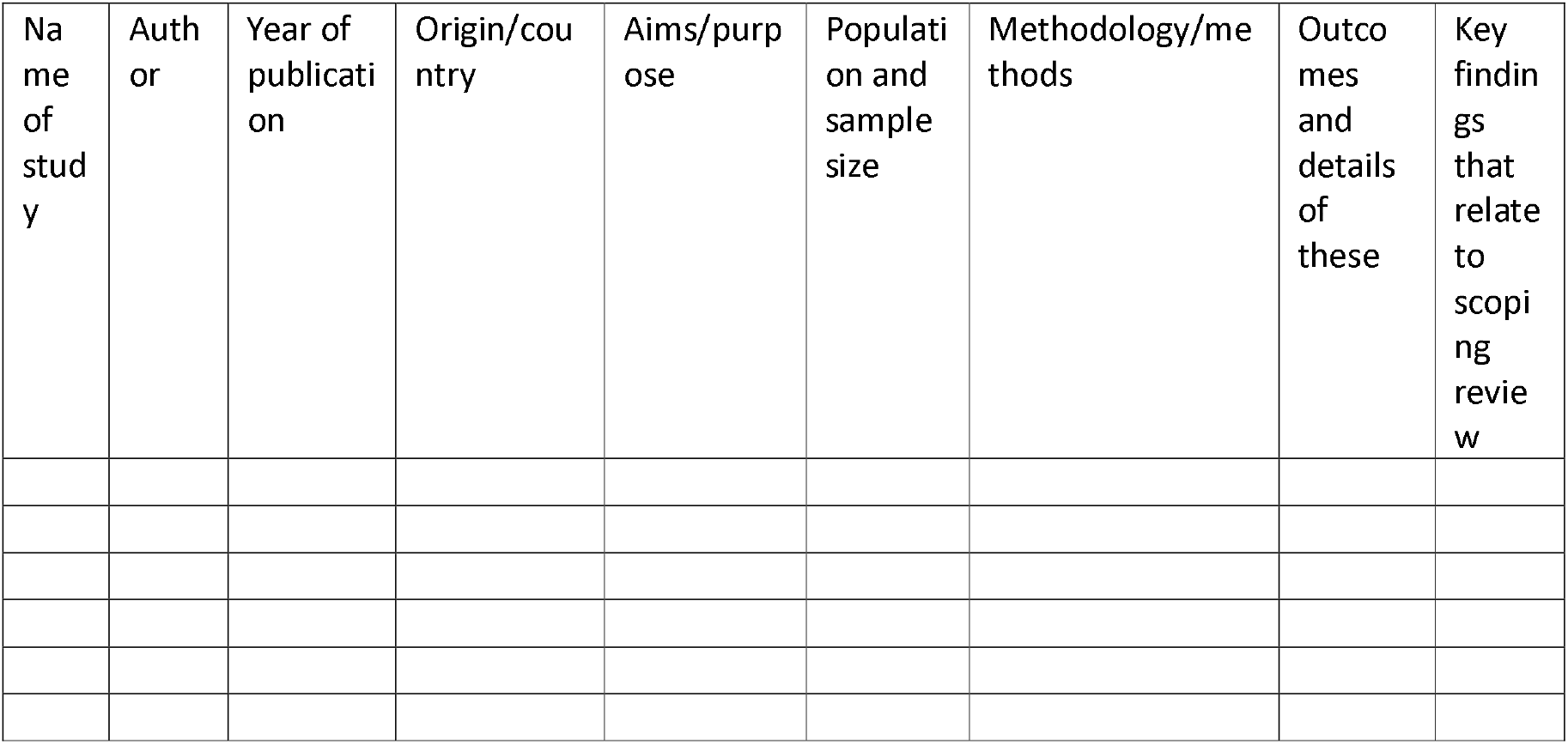
Data extraction instrument

The draft data extraction chart will be modified and revised as necessary during the process of extracting data from each included evidence source.

#### Data analysis and presentation

The intention of the scoping review is to provide a map and summary of available evidence, not to synthesise results into a set of final estimates will findings to inform decision making. The data gathered from the included studies will be discussed in a descriptive text.

## Data Availability

All data produced in the present work are contained in the manuscript

## Ethics and dissemination

This review does not require ethics approval. Our dissemination strategy includes peer review publication, presentation at conferences and relevant stakeholders.

## Details of contributors, and the name of the guarantor

IP developed the original idea for the scoping review as part of a wider research project. IP is responsible for the literature search and the writing of the article. IP led the design of the protocol and methodology and wrote the first draft of the manuscript. DW and JB provided inputs to the methods, designed the search strategy and critically revised the manuscript. DW and JB provided input on the design of the data extraction form. All authors provided valuable inputs to the research questions and subject matter. IP will act as guarantor.

The guarantor accepts full responsibility for the work and/or the conduct of the study and controlled the decision to publish. The corresponding author attests that all listed authors meet authorship criteria and that no others meeting the criteria have been omitted.

## References

1. Saeedi P, Petersohn I, Salpea P, Malanda B, Karuranga S, Unwin N, et al. Global and regional diabetes prevalence estimates for 2019 and projections for 2030 and 2045: Results from the International Diabetes Federation Diabetes Atlas, 9th edition. Diabetes Res Clin Pract [Internet]. 2019;15:107843. Available from: 10.1016/j.diabres.2019.107843

2. Statista. Leading 10 drugs used in the treatment of diabetes in England in 2022, by number of items [Internet]. 2023. Available from: https://www.statista.com/statistics/377865/top-ten-drugs-used-in-diabetes-by-items-in-england/

3. Sharma M, Nazareth I, Petersen I. Trends in incidence, prevalence and prescribing in type 2 diabetes mellitus between 2000 and 2013 in primary care: A retrospective cohort study. BMJ Open [Internet]. 2016;6(1). Available from: 10.1136/bmjopen-2015-010210

4. Alhaji JH. Vitamin B12 deficiency in patients with diabetes on metformin: Arab countries. Nutrients [Internet]. 2022;14(10):2046. Available from: 10.3390/nu14102046

5. Tomkin GH, Hadden DR, Weaver JA, Montgomery DAD. Vitamin-B12 status of patients on long-term metformin therapy. BMJ [Internet]. 1971;2(5763):685–7. Available from: 10.1136/bmj.2.5763.685

6. National Institute for Clinical Excellence (NICE). Type 2 diabetes in adults: Management [Internet]. 2022. Available from: https://www.nice.org.uk/guidance/ng28

7. de Jager J, Kooy A, Lehert P, Wulffel M, van der Kolk J, Bets D. Long-term treatment with metformin in patients with type 2 diabetes and risk of vitamin B-12 deficiency: Randomised placebo-controlled trial. BMJ. 2010;340:c2181. Available from: 10.1136/bmj.c2181

8. Berchtold P, Bolli P, Arbenz U, Keiser G. Intestinale Absorptionsstörung infolge Metforminbehandlung (Zur Frage der Wirkungsweise der Biguanide) [Disturbance of intestinal absorption following metformin therapy (observations on the mode of action of biguanides)]. Diabetologia. 1969;5(6):405–12. German. Available from:10.1007/BF00427979

9. American Diabetes Association (ADA). Classification and diagnosis of diabetes: Standards of medical care in diabetes. Diabetes Care. 2021;44(Suppl 1):S15–S33. Available from: 10.2337/dc21-S002

10. Infante M, Leoni M, Caprio M, Fabbri A. Long-term metformin therapy and vitamin B12 deficiency: An association to bear in mind. World J Diabetes [Internet]. 2021;12(7):916–31. Available from: 10.4239/wjd.v12.i7.916

11. Medicines and Healthcare products Regulatory Authority (MHRA). Metformin and reduced vitamin B12 levels: New advice for monitoring patients at risk [Internet]. 2022. Available from: https://www.gov.uk/drug-safety-update/metformin-and-reduced-vitamin-b12-levels-new-advice-for-monitoring-patients-at-risk

12. Perry LT, Bhasale A, Fabbri A, Lexchin J, Puil L, Joarder M, et al. Comparative analysis of medicines safety advisories released by Australia, Canada, the United States, and the United Kingdom. JAMA Intern Med [Internet]. 2019;179(7):982. Available from: 10.1001/jamainternmed.2019.0294

13. Clinical Knowledge Summaries. Diabetes - type 2: Metformin [Internet]. 2023. Available from: https://cks.nice.org.uk/topics/diabetes-type-2/prescribing-information/metformin/#:~:text=The%20risk%20of%20low%20vitamin,disorders%20such%20as%20Crohn’s%20disease

14. Glasziou P, Haynes B. The paths from research to improved health outcomes. Evid Based Nurs [Internet]. 2005;8(2):36–8. Available from: 10.1136/ebn.8.2.36

15. Zullig LL, Deschodt M, Liska J, Bosworth HB, De Geest S. Moving from the trial to the real world: Improving medication adherence using insights of implementation science. Annu Rev Pharmacol Toxicol [Internet]. 2019;59(1):423–45. Available from: 10.1146/annurev-pharmtox-010818-021348

16. National Institute for Health and Care Research (NIHR). Implementation science [Internet]. 2022. Available from: https://arc-sl.nihr.ac.uk/research-and-implementation/our-research-methods/implementation-science

17. Peters MDJ, Godfrey CM, Khalil H, McInerney P, Parker D, Soares CB. Guidance for conducting systematic scoping reviews. Int J Evid Based Healthc [Internet]. 2015;13(3):141– 6. Available from: 10.1097/xeb.0000000000000050

18. Colquhoun HL, Levac D, O’Brien KK, Straus S, Tricco AC, Perrier L, et al. Scoping reviews: Time for clarity in definition, methods, and reporting. J Clin Epidemiol [Internet]. 2014;67(12):1291–4. Available from: 10.1016/j.jclinepi.2014.03.013

19. Munn Z, Peters MDJ, Stern C, Tufanaru C, McArthur A, Aromataris E. Systematic review or scoping review? Guidance for authors when choosing between a systematic or scoping review approach. BMC Med Res Methodol [Internet]. 2018;18(1). Available from: 10.1186/s12874-018-0611-x

20. Joanna Briggs Institute. Checklist for systematic reviews and research syntheses [Internet]. 2017. Available from: https://joannabriggs.org/ebp/critical_appraisal_tools

21. Arksey H, O’Malley L. Scoping studies: Towards a methodological framework. Int J Soc Res Methodol [Internet]. 2005;8(1):19–32. Available from: 10.1080/1364557032000119616

22. Tricco AC, Lillie E, Zarin W, et al. PRISMA extension for Scoping reviews (PRISMA-SCR): checklist and explanation. Ann Intern Med 2018;169:467–73. Available from: doi:10.7326/M18-0850

23. Page MJ, McKenzie JE, Bossuyt PM, Boutron I, Hoffmann TC, Mulrow CD, et al. The PRISMA 2020 statement: An updated guideline for reporting systematic reviews. BMJ. 2021;372:n71. Available from: 10.1136/bmj.n71

